# Estimation of the final size of the COVID-19 epidemic

**DOI:** 10.1101/2020.02.16.20023606

**Authors:** Milan Batista

## Abstract

In this short paper, the logistic growth model and classic susceptible-infected-recovered dynamic model are used to estimate the final size of the coronavirus epidemic.

## 1 Introduction

One of the common questions regarding an epidemic is its final size. To answer this question various models are used: analytical (Danby 1985, Brauer 2019a, b, Murray 2002), stochastic (Miller 2012), and phenomenological (Fisman D 2014, Pell et al. 2018).

In this note, we attempt to estimate the final epidemic size using the phenomenological logistic growth model (Pell et al. 2018, Chowell G 2014) and the classic susceptible-infected-recovered (SIR) model (Hethcote 2000). With both the models, we obtain a series of daily predictions. The final sizes are then predicted using iterated Shanks transformation (Shanks 1955, Bender and Orszag 1999). The data used for the calculations are taken from *worldmeters* ^1^.

Before proceeding, we note that the final size of the epidemic in its early stage was discussed by Wu et al. (Wu, Leung, and Leung 2020) using the susceptible-exposed-infected-resistant model, by Xiong and Yan (Xiong and Yan 2020) using the exposed-infected-resistant model, by Nesteruk (Nesteruk 2020) using the SIR model, and by Anastassopoulou et al. (Anastassopoulou et al. 2020) using the SIR/death model. These early predictions range from 65000 to a million cases. Roosa et al recently gave short-term forecasts of the epidemic (Roosa et al. 2020).

## 2 Logistic growth model

The logistic growth model originates from population dynamics (Haberman 1998). The underlying assumption of the model is that the rate of change in the number of new cases per capita linearly decreases with the number of cases. Hence, if *C* is the number of cases, and *t* is the time, then the model is expressed as

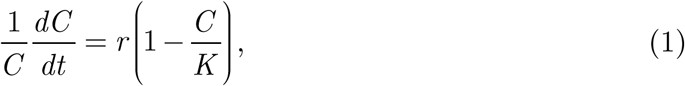

where *r* is infection rate, and *K is* the final epidemic size. If *C* (0) = *C* _0_ is the initial number of cases, then the solution of (1) is

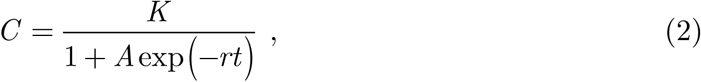

where 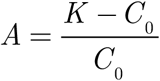. The growth rate, 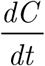, reaches its maximum when 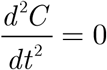. From this condition, we obtain that the growth rate peaks at time t_p._

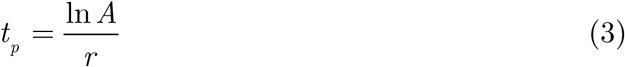

At this time, the number of cases and growth rate are

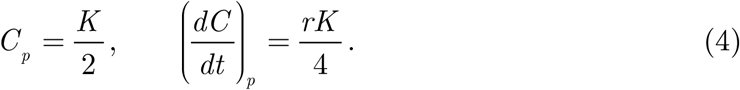

Now, if *C*_1_, *C*_2_, …,*C*_n_ are the number of cases at times *t*_1_, *t*_2_, …,*t*_n_, then the final size predictions of the epidemic based on these data are *K*_1_, *K*_2_, …, *K*_n_. By using Shanks transformation, the predicted final epidemic size is

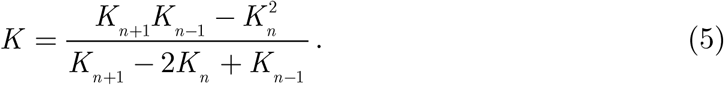

For the practical calculation of the parameters *K* and *r*, we use the MATLAB functions *lsqcurvefit* and *fitnlm*.

## 3 SIR model

The model equations are

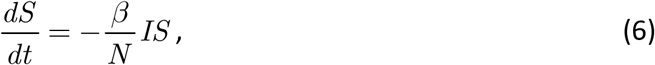

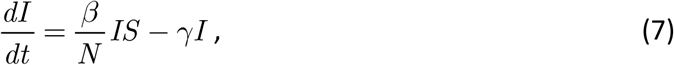

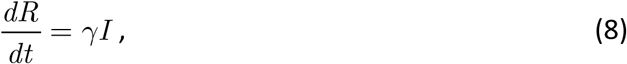

where *t* is time, *S* (*t*) is the number of susceptible persons at time *t, I* = *I* (*t*) is the number of infected persons at time *t, R* (*t*) is the number of recovered persons in time *t, β* is the contact rate, and 1 *ϒ* is the average infectious period. From (1), (2), and (3) we obtain the total population size, *N*.

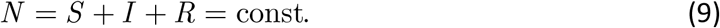

The initial conditions are *S* (0) = *S*_0_, *I* (0) = *I* _0_, and *R* (0) = *R*_0_.

Eliminating *I* from (1) and (3) yields

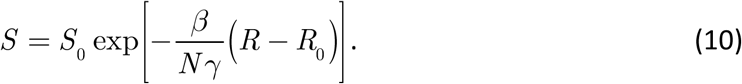

In the limit *t* → ∞, the number of susceptible people left, *S*_∞_, is

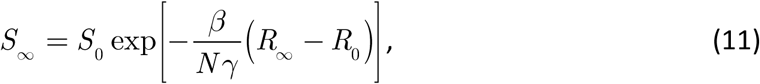

where *R*_∞_is the final number of recovered persons. As the final number of infected people is zero, we have, using (4),

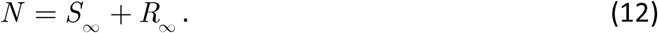

From this and (6), the equation for *R*_∞_is

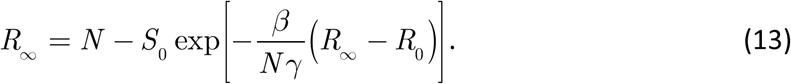

To use the model, we must estimate the model parameters *β, ϒ*, and the initial values *S*_0_ and *I*_0_ from the available data (we set *R* = 0 and *I*_0_ *= C*_0_).

Now the available data is a time series of the total number of cases *C*, i.e.,

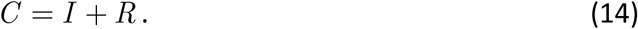

We can estimate the parameters and initial values by minimizing the difference between the actual and predicted number of cases, i.e., by minimizing

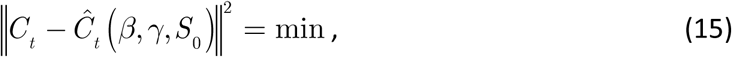

Where *C*_*t*_ = (*C*_1_,*C*_2_,…,*C*_*n*_) are the number of cases at times *t*_1_, *t*_2_, …,*t*_n_ and 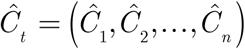 are the corresponding estimates calculated by the model. For practical calculation, we use the MATLAB function `fminsearch`. For the integration of the model equation, we use the MATLAB function *ode45*.

With a series of predicted final number of recovered persons, *R*_∞,1_, *R*_∞,2_,…, *R*_∞,*n*_, we can estimate the series limit by Shanks transformation.

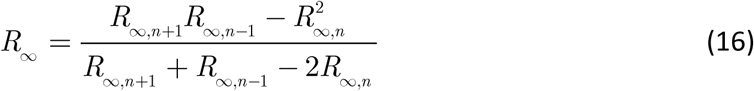

## 4 Results

The results of logistic regression and the SIR model simulation are given in Tables 1 and 2, respectively. The comparison of the predicted final sizes is shown in the graph in Figure 1. We see that both methods converge and with more data, the discrepancy between the predicted values becomes less than 5%. From Table 1, we see that the peak of the epidemic was probably on 9 Feb, 2020.

**Table 1.**
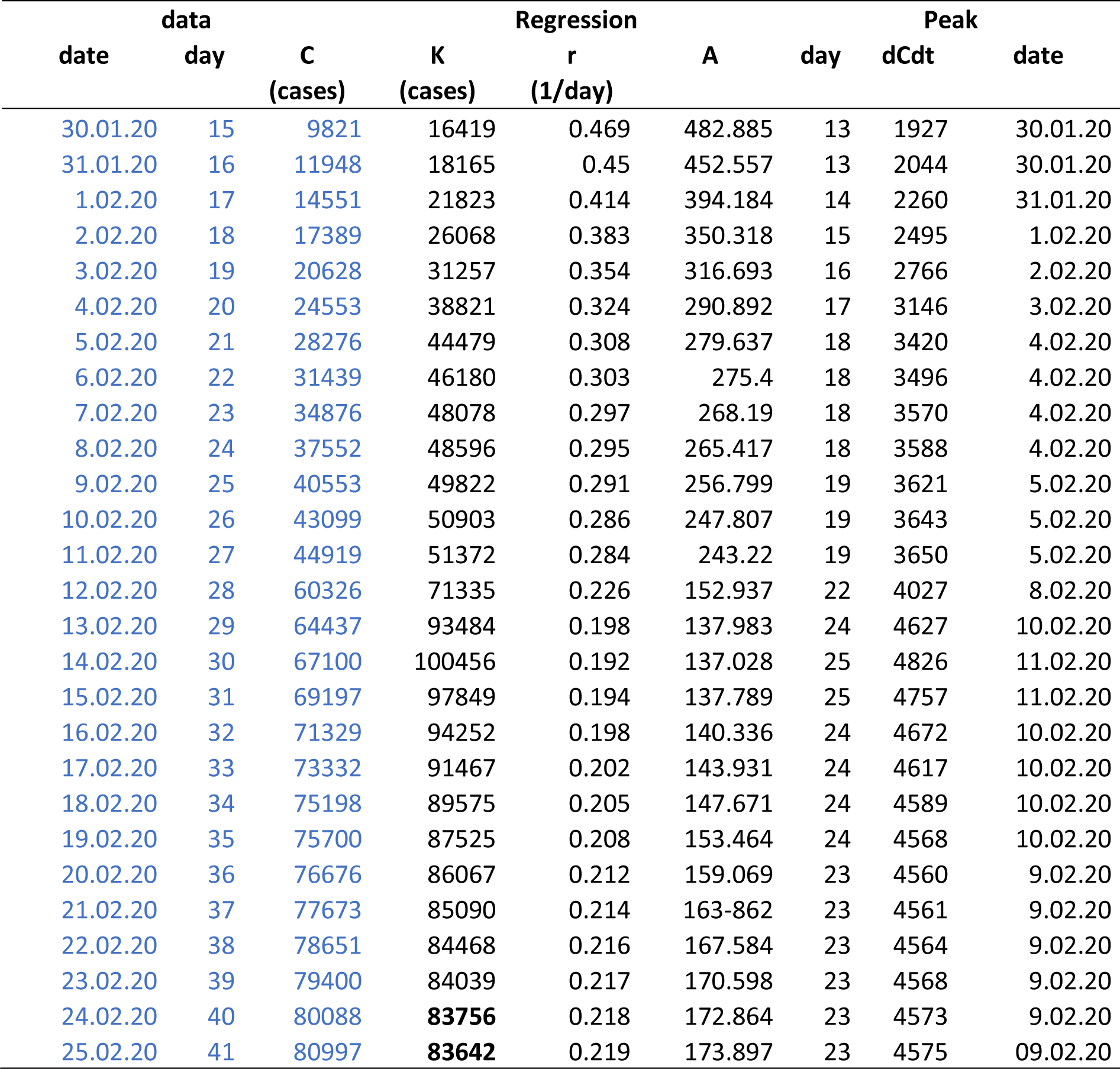
Data and results of logistic regression (see Eqs. (2), (3), (4))

**Table 2.** Results of SIR simulations. After day 28, the method of data collection changes.

| | Day | $N$ | $S_{\infty}$ | $R_{\infty}$ | $R_{\infty,n}/R_{\infty,n-1}$ | $\beta$ | $\gamma$ | $\beta/\gamma$ | $R^2$ |
| --- | --- | --- | --- | --- | --- | --- | --- | --- | --- |
| 12.02.20 | 28 | 551513 | 473888 | 77625 | 1.429 | 2.897 | 2.689 | 1.077 | 0.988 |
| 13.02.20 | 29 | 1300538 | 1189616 | 110922 | 1.048 | 4.026 | 3.854 | 1.045 | 0.988 |
| 14.02.20 | 30 | 1434310 | 1318035 | 116275 | 0.925 | 4.157 | 3.988 | 1.042 | 0.990 |
| 15.02.20 | 31 | 1203132 | 1095609 | 107523 | 0.932 | 3.905 | 3.730 | 1.047 | 0.991 |
| 16.02.20 | 32 | 1002252 | 901990 | 100262 | 0.953 | 3.624 | 3.441 | 1.053 | 0.992 |
| 17.02.20 | 33 | 864976 | 769448 | 95528 | 0.969 | 3.387 | 3.198 | 1.059 | 0.993 |
| 18.02.20 | 34 | 774076 | 681530 | 92546 | 0.969 | 3.205 | 3.010 | 1.065 | 0.994 |
| 19.02.20 | 35 | 683791 | 594070 | 89722 | 0.978 | 2.998 | 2.798 | 1.071 | 0.994 |
| 20.02.20 | 36 | 619431 | 531645 | 87787 | 0.985 | 2.835 | 2.630 | 1.078 | 0.994 |
| 21.02.20 | 37 | 574963 | 488460 | 86503 | 0.990 | 2.712 | 2.504 | 1.083 | 0.994 |
| 22.02.20 | 38 | 544793 | 459126 | 85667 | 0.993 | 2.624 | 2.413 | 1.087 | 0.995 |
| 23.02.20 | 39 | 522591 | 437513 | 85078 | 0.993 | 2.557 | 2.343 | 1.091 | 0.995 |
| 24.02.20 | 40 | 506492 | 421823 | 84669 | 0.995 | 2.506 | 2.291 | 1.094 | 0.995 |
| 24.02.20 | 41 | 497719 | 413262 | 84456 | 0.998 | 2.477 | 2.262 | 1.095 | 0.996 |

**Figure 1.**
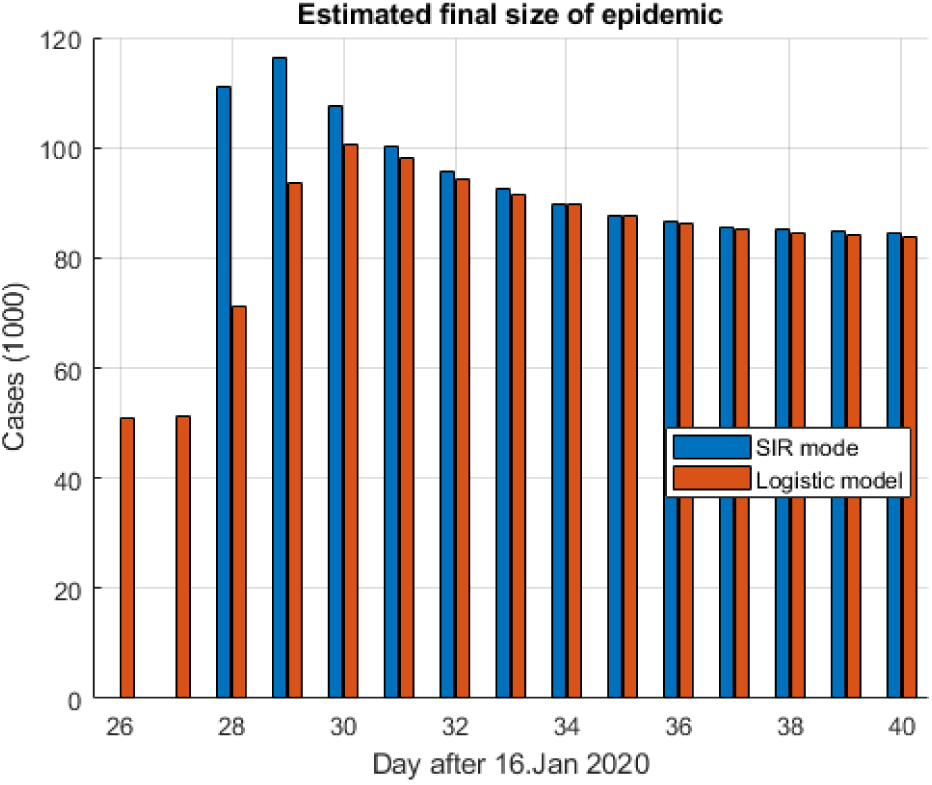
Evaluation of estimated final size of coronavirus epidemic (data until 20 Feb, 2020)

In Figure 2, the time evaluation of the cases is shown, where we can see a good agreement between the models and the actual data. From Table 3, we see that the logistic regression model has a high coefficient of determination of 0.996, while the p-value (< 0.000) indicates that all the regression parameters are statistically significant.

**Table 3.** Estimated logistic model parameters for data until 25 Feb, 2020

|  | Estimate | SE | tStat | pValue |
| --- | --- | --- | --- | --- |
| K | 83643 | 1239.9 | 67.46 | 3.5853e-41 |
| r | 0.21882 | 0.0083521 | 26.199 | 6.2629e-26 |
| A | 173.89 | 29.73 | 5.8492 | 9.1682e-07 |

Number of observations: 41, Error degrees of freedom: 38
Root Mean Squared Error: 2.15e+03
R-Squared: 0.996, Adjusted R-Squared 0.995
F-statistic vs. zero model: 6.35e+03, p-value = 2.42e-51

**Table 3.** Iterated Shanks transformation for logistic model

| day | $K$ | $K(K)$ | $K(K(K))$ | $K(K(K(K)))$ |
| --- | --- | --- | --- | --- |
| 35 | 87524 |  |  |  |
| 36 | 86067 | 83101 |  |  |
| 37 | 85090 | 83386 | 83236 |  |
| 38 | 84469 | 83071 | 83372 | 83231 |
| 39 | 84039 | 90072 | 86718 |  |
| 40 | 83576 | 83634 |  |  |
| 41 | 83642 |  |  |  |

**Figure 2.**
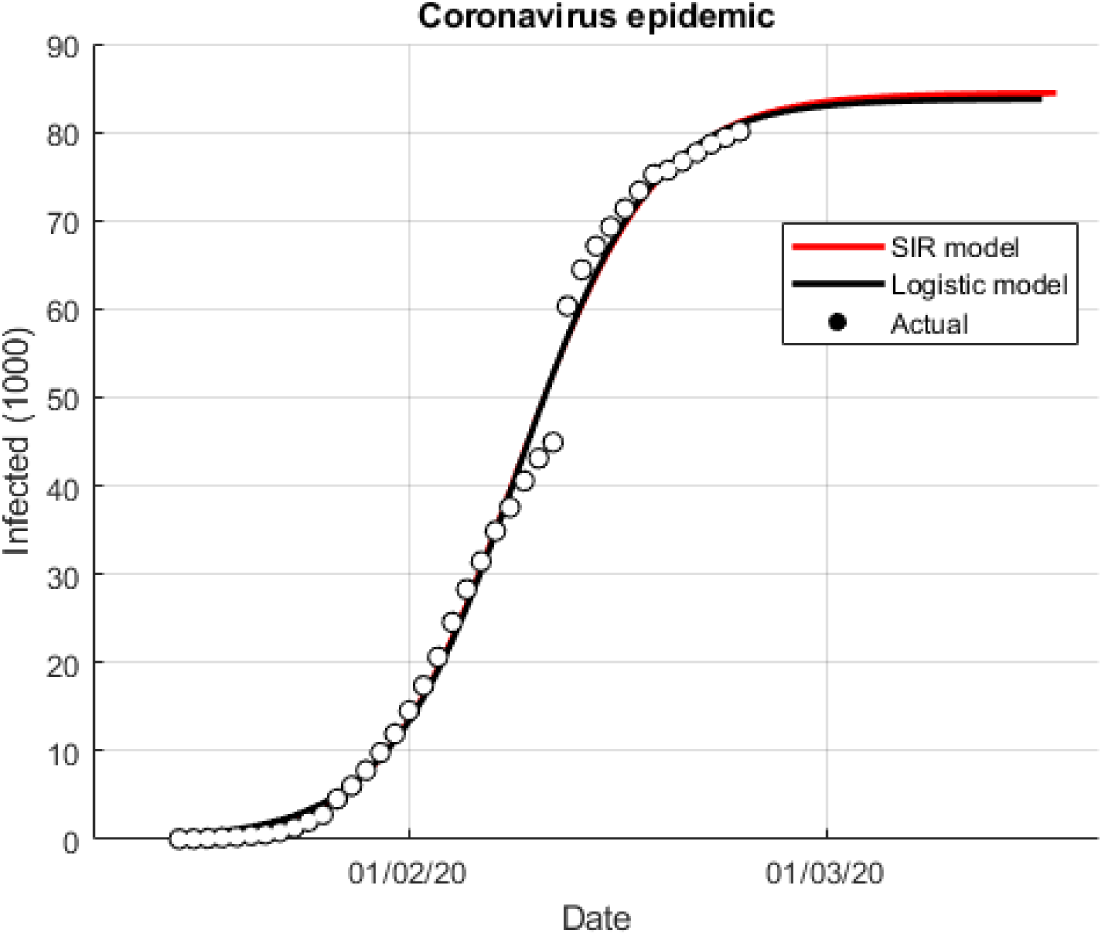
Predicted evaluation of coronavirus epidemic (data until 20 Feb, 2020)

In Tables 3 and 4, the iterated Shanks transformations for the predicted series of the final epidemic size are given. It appears that the predictions of the logistic model tend to the final size of 83231 cases, while the SIR model predictions converge to 83640 cases. Thus, the discrepancy is less than 0.5%.

**Table 4.**
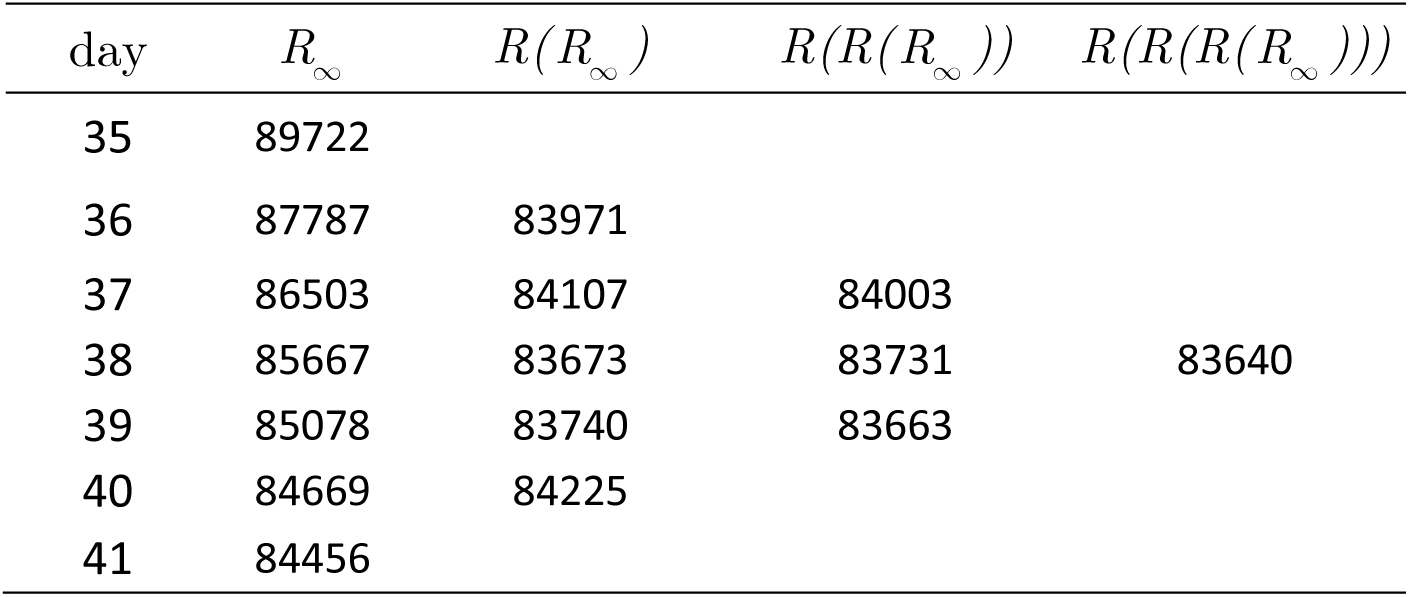
Iterated Shanks transformation for SIR model

## 5 Short term forecasting

The models used are data-driven, so they are as reliable as data are. Namely, as can be seen from the graph in Figure 2 at the beginning, we have exponential growth. Then until 11 Feb, one can predict the final epidemic size of about 55000 cases. However, the collection of data changes and we have a jump of about 15000 new cases on 12. Feb. On 20 Feb we have another change in trend; the data begin to shows almost linear trend (See Fig 3). While the above models show that the epidemic is slow down, the linear trend predicts about 873 new cases per day (see Table 5).

**Table 5.** Short term forecasting with the logistic and linear model. The linear model predicts 873 new cases per day.

| Date | Day | Actual | Logistic<br>model | Error<br>% | Linear<br>model | Error<br>% | Actual<br>cases/day | Predicted<br>cases/day |
| --- | --- | --- | --- | --- | --- | --- | --- | --- |
| 20.02.20 | 35 | 75700 | 75890 | 2.111 | 75837 | 0.180 |  |  |
| 21.02.20 | 36 | 76676 | 77298 | 2.337 | 76709 | 0.044 | 976 | 1408 |
| 22.02.20 | 37 | 77673 | 78468 | 2.267 | 77582 | 0.117 | 997 | 1170 |
| 23.02.20 | 38 | 78651 | 79434 | 2.004 | 78455 | 0.249 | 978 | 966 |
| 24.02.20 | 39 | 79400 | 80227 | 1.859 | 79328 | 0.090 | 749 | 793 |
| 25.02.20 | 40 | 80088 | 80876 | 1.644 | 80201 | 0.141 | 688 | 649 |
| 26.02.20 | 41 | 80997 | 81405 | 1.036 | 81074 | 0.095 | 909 | 529 |
| 27.02.20 | 42 |  | 81836 |  | 81947 |  |  | 431 |
| 28.02.20 | 43 |  | 82185 |  | 82820 |  |  | 349 |
| 29.02.20 | 44 |  | 82467 |  | 83693 |  |  | 282 |
| 1.03.20 | 45 |  | 82696 |  | 84566 |  |  | 229 |
| 2.03.20 | 46 |  | 82880 |  | 85439 |  |  | 184 |
| 3.03.20 | 47 |  | 83029 |  | 86312 |  |  | 149 |

**Figure 3.**
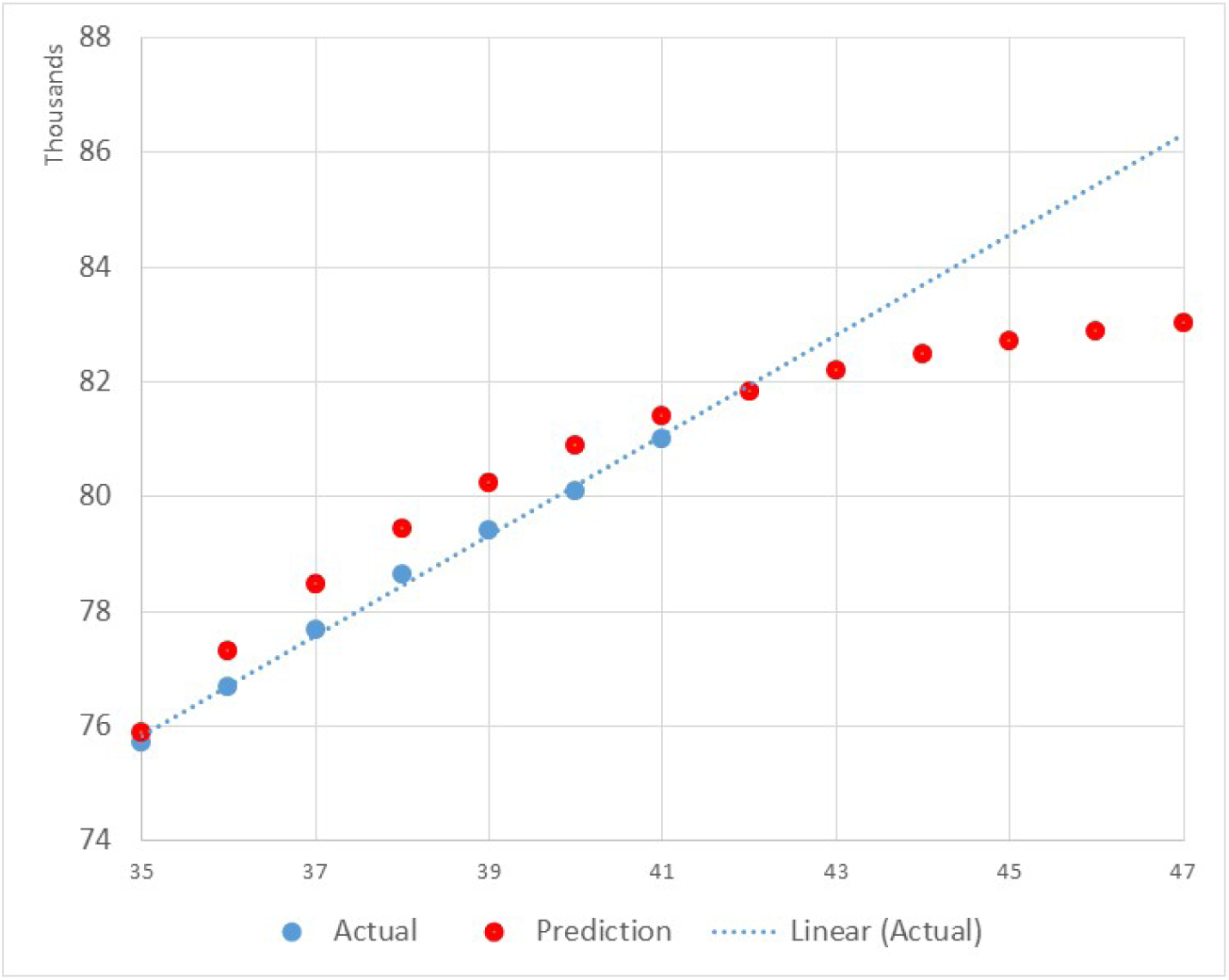
Short-term forecasting from 20 Feb 2020.

## 6 Conclusion

On the basis of the available data, we can now predict that the final size of the coronavirus epidemic using the logistic model will be approximately 83700 (±1300)?cases and that the peak of the epidemic was on 9 Feb 2020. A more optimistic final size of 83300 cases is obtained using the Shanks transformation. Similar figures are obtained using the SIR model, where the predicted size of the epidemic is approximately 84500, and the Shanks transformation lowers this number to about 83700 cases. Naturally, the degree of accuracy of these estimates remains to be seen.

In conclusion, qualitatively, both models show that the epidemic is moderating, but recent data show a linear upward trend. The next few days will, therefore, indicate in which direction the epidemic is heading.

PS. Today it is more or less clear that the predictions of the article apply only to China. By February 20, 99% of the case was from China. The linear trend in data from Feb 20 onward meant a decreasing number of infected in China and increasing infected elsewhere in the world. In other words, in China, the epidemic is slowing down, however, it is now developing elsewhere in the world. We note that the forecasting methods used in this article are inapplicable in the early stages of an epidemic.

## Data Availability

The data used are from
https://www.worldometers.info/coronavirus/

## Footnotes

1 https://www.worldometers.info/coronavirus/

